# Anti-CD38 therapy impairs SARS-CoV-2 vaccine response in multiple myeloma patients

**DOI:** 10.1101/2021.08.08.21261769

**Authors:** S. Henriquez, J. Zerbit, T. Bruel, A. Ouedrani, D. Planas, P. Deschamps, I. Staropoli, J. Hadjadj, B. Varet, F. Suarez, N. Ermark, D. Bouscary, L. Willems, G. Fouquet, J. Decroocq, P. Franchi, B. Deau-Fischer, B. Terrier, J. Tamburini, L. Chatenoud, O. Schwartz, M. Vignon

**Author notes:** **Address correspondence:** Dr. Marguerite Vignon, Department of Clinical Hematology, Hôpital Cochin, 27, rue du Faubourg Saint-Jacques, 75014 Paris, France. These authors contributed equally to this work (S. Henriquez, J. Zerbit, T. Bruel, A. Ouedrani).

## Abstract

Multiple myeloma (MM) patients are at risk of fatal outcome after SARS-CoV-2 infection. Preliminary data suggest that MM patients have an impaired response to vaccination. This prospective study analyzed the humoral and cellular immune responses to two doses of BNT162b2 in 72 MM patients, including 48 receiving anti-CD38 immunotherapy. Results evidenced that MM patients display lower levels of SARS-CoV-2 specific IgG and IgA antibodies and decreased neutralization of alpha and delta variants when compared to healthy controls. They also showed decreased numbers of circulating IFNγ-producing Spike SARS-CoV-2 specific T lymphocytes. This defective immune response was particularly marked in patients receiving anti-CD38 immunotherapy. Furthermore, a retrospective investigation of MM patients among COVID-19-related death in the Paris area suggested a limited efficacy of BNT162b2 in patients treated with anti-CD38. Overall, these results show a decreased immunogenicity of BNT162b2 in MM patients and stress the need for novel strategies to improve SARS-CoV-2 prophylaxis in immunocompromised individuals.

## Introduction

Patients with multiple myeloma (MM) have an increased risk of SARS-CoV-2 complications and death compared to the general population, particularly in case of progressive disease (PD) status, older age (>60 years old) and renal impairment (Chari *et al*., 2020; Dufour *et al*., 2020). Accordingly, MM patients were considered as a high-priority population for vaccination against SARS-CoV-2.

Recent clinical trials with mRNA vaccines did not include immunocompromised patients. Suboptimal vaccination responses against a range of pathogens is broadly reported in MM patients under treatment (Robertson *et al*., 2000), but data describing the immunogenicity of SARS-CoV-2 vaccines in MM patients are scarce (Polack *et al*., 2020; Baden *et al*., 2021). Preliminary results suggest an impaired immune response to a single SARS-CoV-2 vaccine dose in MM patients regardless their therapeutic regimen (Terpos *et al*., 2021). How MM patients respond to 2 doses of vaccine remains to be determined.

Immunotherapies with monoclonal anti-CD38 antibodies are increasingly used in MM patients and were recently approved in first line in combination with other agents (immunomodulatory drugs -IMIDs or proteasome inhibitors -PI) and high-dose steroids (Moreau *et al*., 2019). These targeted immunotherapeutic approaches could impair the vaccine response, as CD38 is expressed by both normal and malignant plasma cells (Flores-Montero *et al*., 2016), similar to the well-documented negative effect of anti-CD20 immunotherapy on vaccine response (Rieger *et al*., 2018; Baker *et al*., 2020). The emergence of SARS-CoV-2 variants with partial immune escape potential represent an additional risk for individuals with sub-optimal vaccine response (Planas, Veyer, *et al*., 2021). Thus, a better understanding of the immune response to SARS-CoV-2 vaccines in MM patients, according to disease status, ongoing therapy and circulating variants of concern is critically needed.

Here, we prospectively investigated the production of anti-Spike immunoglobulins (IgG and IgA) as well as neutralizing antibodies against alpha (B.1.1.7) and delta (B.1.617.2) variants in 72 MM patients and 20 control individuals having received a full vaccination regimen (i.e 2 doses) with a mRNA vaccine (Pfizer-BioNTech BNT162b2) at a single tertiary-care center. We also assessed in 32 patients the specific anti-SARS-CoV-2 T cell response using EliSpot. Finally, we reported SARS-CoV-2 mortality in MM receiving or not anti-CD38 from January 2020 to July 2021 at Assistance Publique – Hôpitaux de Paris (AP-HP). Our study shows an impaired immune response to SARS-CoV-2 mRNA vaccine in MM patients, particularly those treated with anti-CD38, with a possible impact on SARS-CoV-2-related mortality.

## Material and methods

### Patients

Patients were prospectively included from January 2021 to June 2021 at Groupe Hospitalo-Universitaire AP-HP. Centre-Université de Paris. Inclusion criteria were: age >18 years, receiving active MM treatment or with treatment discontinued for less than one year, and having received two doses of mRNA Pfizer-BioNTech BNT162b2 vaccine 28 days apart. Treatment indication for MM followed IMWG recommendations (Rajkumar *et al*., 2014). Exclusion criteria were opposition to data collection or inability to give consents. A written informed consent was obtained from all the patients for clinical data recording and blood samples collection in agreement with the declaration of Helsinki. The study protocol was approved by the institutional review board of Cochin Hospital (CLEP N°: AAA-2021-08041). Data collection has been declared to the National Commission for Data Processing and Freedoms. The study was registered with the French National Agency for Medicines and Health Products Safety (number 2021-A02124-37).

Health caregiver from the same hospital constituted the control cohort. They received two vaccine injections following the same modalities as for MM patients, and history of SARS-CoV-2 infection before or after vaccination was screened. They also gave written informed consent. Control cohort was previously described (Hadjadj *et al*., 2021, submitted).

### Study procedure

Blood samples from patients and controls were collected the day before vaccination (referred to as T0 for time 0), at 28 days before the second dose (referred to as M1 for month 1) and one to two months after completion of vaccination (referred to as M3) to investigate humoral response to SARS-CoV-2 vaccine. Whole blood specimens were collected in a subset of patients at M3 to perform enzyme-linked immunospot (EliSpot) assays on peripheral blood mononuclear cells to investigate the cellular immune response.

Clinical data were recorded and included demography, MM history (evolution time, number of lines including autologous stem cell transplantation (ASCT) and anti-CD38, response to treatment at the time of inclusion), previous history of SARS-CoV-2 infection, and biological parameters including gammaglobulin levels and lymphocyte count at the time of vaccination. The occurrence of SARS-CoV-2 infection after vaccination was recorded. Patients were considered as having progressive disease if a therapeutic change was done at least three months before or any time after inclusion. High-dose steroid treatment was defined as administration of at least 20mg dexamethasone weekly.

### S-Flow assay

Anti-spike IgG and IgA antibodies were quantified using the S-Flow assay as previously described (Grzelak *et al*., 2020). Briefly, 293T cells stably expressing the S protein (293T Spike cells) and 293T Empty cells as control were incubated with sera (1:300 dilution) in PBS containing 0.5% BSA and 2 mM EDTA. After 30min of incubation at 4°C, cells were washed with PBS, and stained using anti-Hu IgG FC-Alexafluor 647 antibody (109-605-170, Jackson ImmunoResearch) and anti-Hu IgA alphaChain-Alexafluor 488 antibodies (109-545-011, Jackson ImmunoResearch) for 30min at 4°C. Cells were then washed with PBS and fixed for 10 min using 4% paraformaldehyde (PFA). Data were acquired on an Attune NxT instrument (Life Technologies). The positivity of a sample was defined as a specific binding above 40% for IgG and 20% for IgA (Grzelak *et al*., 2020). To standardize the results, a binding units (BU) was calculated using a serially diluted human anti-S monoclonal antibody as reference (Grzelak *et al*., 2021). The logarithm of the median of fluorescence of each sample was reported on the curve to obtain an equivalent value (in ng/mL) of antibody concentration in logarithm.

### Virus strains

The Alpha variant (B.1.1.7) originated from an individual in Tours (France) returning from United Kingdom. The Delta variant (B.1.617.2) originated from a nasopharyngeal swab of a hospitalized patient returning from India. Both patients provided informed consent for the use of the biological materials. The variant strains were isolated from nasal swabs using Vero E6 cells and amplified by one or two passages. Titration of viral stocks was performed on Vero E6 cells, with a limiting dilution technique allowing a calculation of the 50% tissue culture infectious dose, or on S-Fuse cells. Viruses were sequenced directly on nasal swabs and after one or two passages on Vero cells.

### S-Fuse neutralization assay

U2OS-ACE2 GFP1–10 or GFP 11 cells, also termed S-Fuse cells, become GFP^+^ cells when they are productively infected with SARS-CoV-2 (Planas, Bruel, *et al*., 2021). Cells were mixed at a 1:1 ratio and 8⍰×⍰10^3^ cells/well were seeded in μClear 96-well plate (Greiner Bio-One). SARS-CoV-2 strains were incubated with sera at the indicated dilutions for 15⍰min at room temperature and added to S-Fuse cells. 18⍰h later, cells were fixed with 2% paraformaldehyde, washed with PBS and stained with Hoechst (1:1,000 dilution; Invitrogen). Images were acquired with an Opera Phenix high-content confocal microscope (PerkinElmer). syncytia and nuclei were counted using the Harmony software (PerkinElmer). The number of syncytia was used to determine neutralization with the formula: 100⍰×⍰(1⍰−⍰(value with serum⍰−⍰value in ‘noninfected’)/(value in ‘no serum’⍰−⍰value in ‘noninfected’)). ED_50_ of neutralization (i.e titers) were calculated using a reconstructed curve of neutralization and reciprocal dilution. Of note, we previously reported a correlation between neutralization titers obtained with the S-Fuse reporter assay and a pseudovirus neutralization assay (Sterlin *et al*., 2021). Cells were tested negative for mycoplasma. All Sera were heat inactivated 30⍰min at 56⍰°C before use. Positivity for antibody neutralization production was defined as above 30.

### T-cell response using enzyme-linked immunoSpot (EliSpot)

Freshly isolated peripheral blood mononuclear cells (PBMCs) were isolated from blood collected in vacutainers containing lithium heparin as anti-coagulant (BD Vacutainer, Le Pont de Claix, France). After density gradient separation of PBMC using Uni-SepMAXI tubes (Eurobio, Courtaboeuf, France) according to the manufacturer’s instructions, lymphocytes were enumerated by flow cytometry using the BD Tritest™ CD3FITC/ CD8PE/ CD45PerCP and BD Trucount™ Tubes (BD Biosciences, Le Pont de Claix, France). The EliSpot assay was adapted from the procedure previously described (Candon et al., 2009; Hueso et al., 2020). Briefly, day 0, sterile PVDF strips (Millipore, Saint-Quentin-en-Yvelines, France) were coated overnight at 4°C with an IFNγ antibody (U-CyTech, Utrecht, Netherlands). Day 1, strips were blocked with culture medium (RPMI-1640, with L-glutamine and sodium bicarbonate (Sigma-Aldrich, Molsheim, France) supplemented with 10% human AB serum) into 37°C humidified incubator, 9% CO2 during 1 hour. PBMCs were then seeded at 200 000 CD3+T cell/well in duplicates and stimulated for 18-20hrs with individual pool of 15-mer peptides with 11 amino acids overlap at a final concentration of 10 μmol/L. Day 2, PBMCs were removed and IFNγ secretion revealed using a biotin-conjugated IFNγ antibody (U-CyTech), streptavidin-horseradish peroxidase (UCyTech) and 3-amino-9-ethylcarbazole (AEC) (U-CyTech). Spots were enumerated using an automated EliSpot reader (Autoimmune Diagnostika (AID) reader, Strassberg, Germany) and systematically checked by an experienced operator. To identify SARS-CoV-2-Spike-specific T cells, we used commercially available pool derived from a peptide scan through SARS-CoV-2 N-terminal fragment (pool S1) and C-terminal fragment (pool S2) (JPT Peptide Technologies GmbH, BioNTech AG, Berlin, Germany). Negative controls were cells in culture medium. Positive controls were phytohemagglutinin PHA-P (Sigma-Aldrich) at a final concentration of 1 mg/ml and the CEFX Ultra SuperStim Pool (JPT Peptide Technologies GmbH, BioNTech AG, Berlin, Germany) at a final concentration of 10 μmol/L. The CEFX Ultra SuperStim Pool, contained peptide epitopes for a broad range of HLA sub-types and different infectious agents. Results were expressed as Spot Forming Unit (SFU)/106 CD3+ T lymphocytes after subtraction of background values from wells with non-stimulated cells. The EliSpot was technically validated when mean number of spots in unstimulated wells was under than or equal to 10. The detection threshold was set at 3SD above the average basal reactivity (mean spot number in RPMI wells), or at least fixed to 11 SFU/106 CD3+ to rule out false positives where background was very low. Relevant clinical positivity thresholds for peptide S1 pool were determined in the lab as following: 20 SFU/10^6^ CD3+. Relevant clinical positivity thresholds for each peptide pool were determined in the lab as following: 20 SFU/10^6^ CD3+ for S1 pool and 60 SFU/10^6^ CD3+ for S2 pool.

### Health care data

Data on COVID 19-related mortality were obtained from the hospital’s electronic health records (HER) using the Informatics for Integrated Biology and the Bedside (i2b2) platform. A cross search using diagnostic codes and treatment reports was performed on all 39 AP-HP hospital’s EHR from January 2020 to July 2021 in order to intersect the COVID-19 deaths, patients with myeloma, and those treated with anti-CD38. Data on the number of anti-CD38 therapies administrated to MM patients at these care centers from January 2020 to July 2021 were obtained from AGEPS (General Agency of Equipments and Health Products). Analysis focused on two epidemic peak periods before (period 1, March-July 2020) and after (period 2, March-July 2021) vaccination program onset.

### Statistical analysis

Statistical analysis of categorical variables was performed using the Fisher exact test. Differences between the mean values between two experimental groups were analyzed using a Mann-Whitney test for non-parametric data. Differences between more than two experimental groups were investigated by one-way analysis of variance (ANOVA) using the non-parametric Kruskal-Wallis test for multiple comparisons. Statistical analyses were performed using Prism software 9.2.0 (GraphPad).

*P<0.05, **P<0.01, ***P<0.001, ****P<0.0001, ns: not significant.

## Results

### Description of patients and control populations

We included 72 consecutive MM patients in our study, among whom 11 had a clinical SARS-CoV-2 infection history more than three month before vaccination. They received the first injection of BNT162b2 (Comirnaty®) from January 18^th^ to March 21^st^, 2021. All patients received the second injection four weeks apart. At the time of vaccination, 48 patients were treated with anti-CD38 immunotherapy-based regimen (anti-CD38 group), and 24 patients were not (non-anti-CD38 group) (**Figure 1**). **Table 1** summarized the characteristics of patients according to the ongoing therapeutic strategy. Patients in the anti-CD38 group received more treatment lines and more patients had an active treatment at inclusion. Anti-CD38 therapy was associated with chemotherapy in 33 cases (69%). Twenty (83%) patients from the non-anti-CD38 group were on-therapy, including immunomodulatory drugs (IMIDs), proteasome inhibitor (PI) or IMIDs+PI combination in 11, 4 and 5 cases, respectively. Gammaglobulins (γ-globulins) level was significantly lower in the anti-CD38 group, and more patients from this group had γ-globulin level <5 g/l. Patients with documented SARS-CoV-2 infection before vaccination were analyzed separately for immune response parameters. We included 20 SARS-CoV-2-naive healthy volunteer subjects in a control cohort. They received two vaccine injections (**Supplemental Figure 1A**).

**Table 1.** Characteristics of MM patients dependent on anti-CD38 immunotherapy. Min: minimum; max: maximum; Ev.: length of MM evolution; y: years; Lines: number of therapeutic lines; ASCT: autologous stem cell transplantation; PD: progressive disease; MGRS: monoclonal gammopathy of renal significance; PI: proteasome inhibitor; Steroids: high-dose steroid therapy ie. more than 20mg dexamethasone per week; γ: corrected gammaglobulin levels; Ig: immunoglobulins; Ly: lymphocytes.

| Variables | All (n=72) | Anti-CD38 (n=48) | Non-anti-CD38 (n=24) | p-value | test |
| --- | --- | --- | --- | --- | --- |
| **Demography** |  |  |  |  |  |
| Age, mean (min-max) | 69.86 (40.8-92.1) | 69.68 (40.8-87) | 70.2 (53-92.1) | 0.68 | t-test |
| Male Gender (%) | 37 (57%) | 28 (58%) | 9 (37.5%) | 0.25 | Fischer |
| **Disease characteristics** |  |  |  |  |  |
| Ev., y, mean (min-max) | 5.69 (0-19) | 6.45 (0-19) | 4.16 (0-14) | 0.09 | t-test |
| Lines, mean (min-max) | 2.43 (1-12) | 2.81 (1-12) | 1.66 (1-7) | **0.0001** | t-test |
| 1st line vs other, n (%) | 26 (37%) | 8 (16.6%) | 18 (75%) | **<0.001** | Fischer |
| ASCT, n (%) | 13 (18%) | 3 (6.25%) | 10 (41.6%) | **0.0006** | Fischer |
| PD, n (%) | 17 (23.6%) | 10 (20.8%) | 7 (29.2%) | 0.55 | Fischer |
| MGRS, n (%) | 8 (11%) | 5 (10.4%) | 3 (12.5%) | 0.99 | Fischer |
| **Therapy status** |  |  |  |  |  |
| On therapy, n (%) | 65 (90.2%) | 48 (100%) | 20 (83%) | **0.003** | Fischer |
| Anti-CD38, n (%) | 48 (66%) | 45 (100%) | 0 (0%) |  | N/A |
| IMiDs, n (%) | 34 (47.2%) | 23 (47.9%) | 11 (45.8%) | 0.99 | Fischer |
| PI, n (%) | 13 (18%) | 9 (18.75) | 4 (16.6%) | 0.99 | Fischer |
| IMiDs+PI, n (%) | 6 (8.3%) | 1 (2%) | 5 (20.8%) | **0.0139** | Fischer |
| Steroid, n (%) | 26 (34%) | 17 (35.4%) | 5 (20.8%) | 0.6 | Fischer |
| **Biological parameters** |  |  |  |  |  |
| $\gamma$ (g/L), mean (min-max) | 4.65 (0.8-11.9) | 3.5 (0.8-11.9) | 5.9 (1.9-10.1) | **0.0011** | t-test |
| $\gamma < 5$ g/l, n (%) | 47 (65.2%) | 37 (77%) | 10 (41.6%) | **0.0042** | Fischer |
| Ig substitution, n (%) | 12 (16.6%) | 8 (18%) | 6 (19%) | 0.99 | Fischer |
| Ly $< 0.8 \times 10^9/l$ n (%) | 24 (33%) | 16 (33%) | 8 (33%) | 0.99 | Fischer |
| **Covid infection** |  |  |  |  |  |
| History, n (%) | 11 (15.2%) | 6 (12.5%) | 5 (20.8%) | 0.32 | Fischer |
| After vaccination, n (%) | 4 (5.5%) | 4 (8.3%) | 0 (0%) | 0.29 | Fischer |

**Figure 1.**
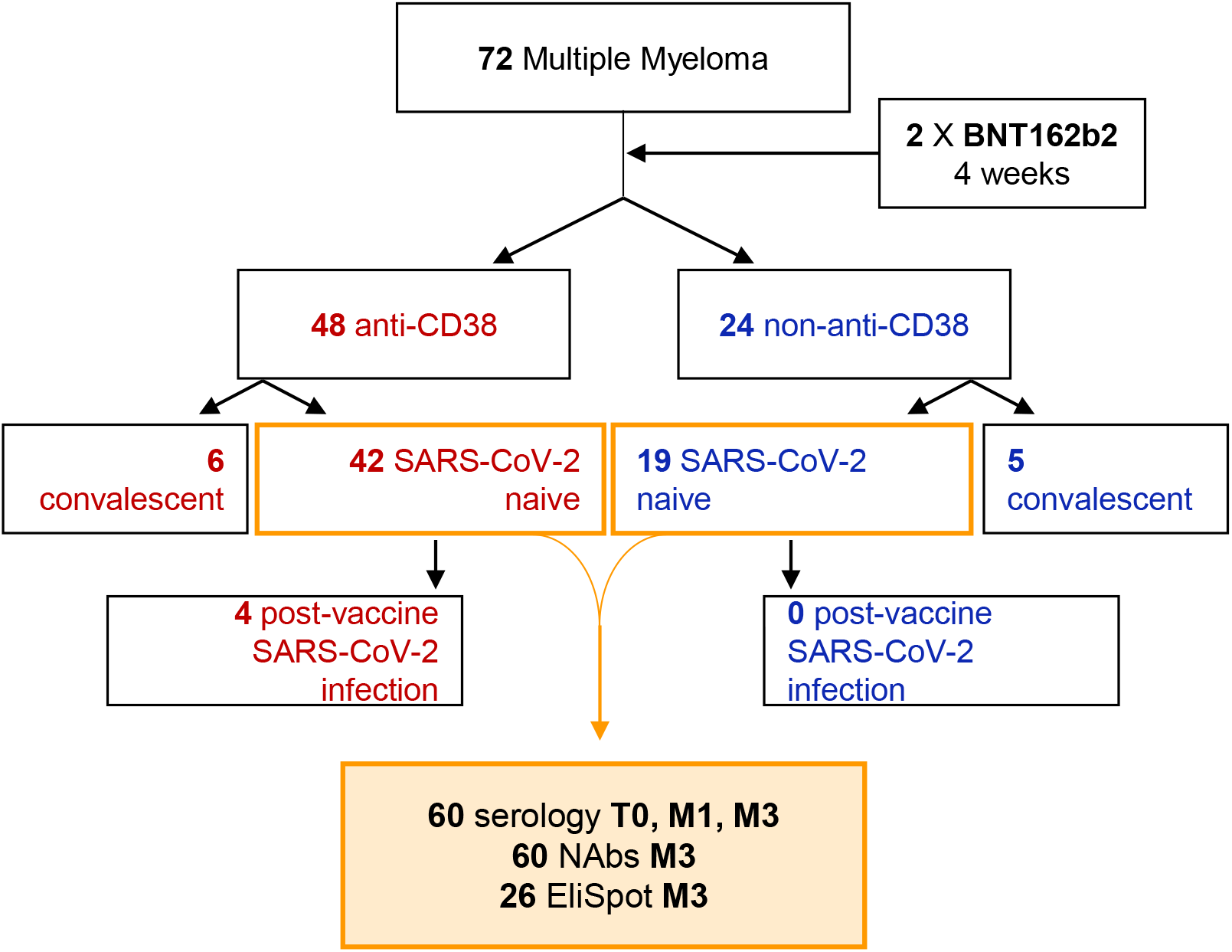
Clinical response to BNT162b2 in multiple myeloma patients. Flow-chart of MM patients prospectively included in our study. 2 X BNT162b2: two doses of BNT162b2 mRNA vaccine. Convalescent: having suffered from SARS-CoV-2 infection more than three month before inclusion. Serology: specific anti-SARS-CoV-2 IgG or IgA quantification by the S-flow method; NAbs: neutralizing antibodies against alpha or delta SARS-CoV-2 variants; EliSpot: IFNγ production in response to S1 or S2 SARS-CoV-2 proteins. M1 and M3: one and three months after the first dose of BNT162b2.

### Impaired humoral response to BNT162b2 in multiple myeloma patients

We investigated the specific anti-SARS-CoV-2 IgG and IgA production using the S-flow technique (referred to as antibody binding or Ab binding, **Supplemental Figure 2A**) before, one (M1) and three (M3) months after the first BNT162b2 injection. In the control group, 100% of subjects achieved IgG seroconversion at M1, after one vaccine dose. In SARS-CoV-2-naive MM patients, only 44% experienced seroconversion at M1, while 85% had IgG seroconversion at M3 (**Figure 2A**). The levels of SARS-CoV-2 IgG and IgA were significantly lower at M1 in MM patients than in controls, and remained lower for IgA at M3 (**Figure 2B**). We then compared the production of SARS-CoV-2 specific IgG and IgA at M3 between the anti-CD38 and non-anti-CD38 groups of MM patients. IgG levels were decreased in the anti-CD38 group. IgA levels were low and similar between the two groups (**Figures 2C-D**). These results suggest that MM patients who are actively treated, particularly with anti-CD38 immunotherapies, had an impaired production of SARS-CoV-2 specific IgG and IgA.

**Figure 2.**
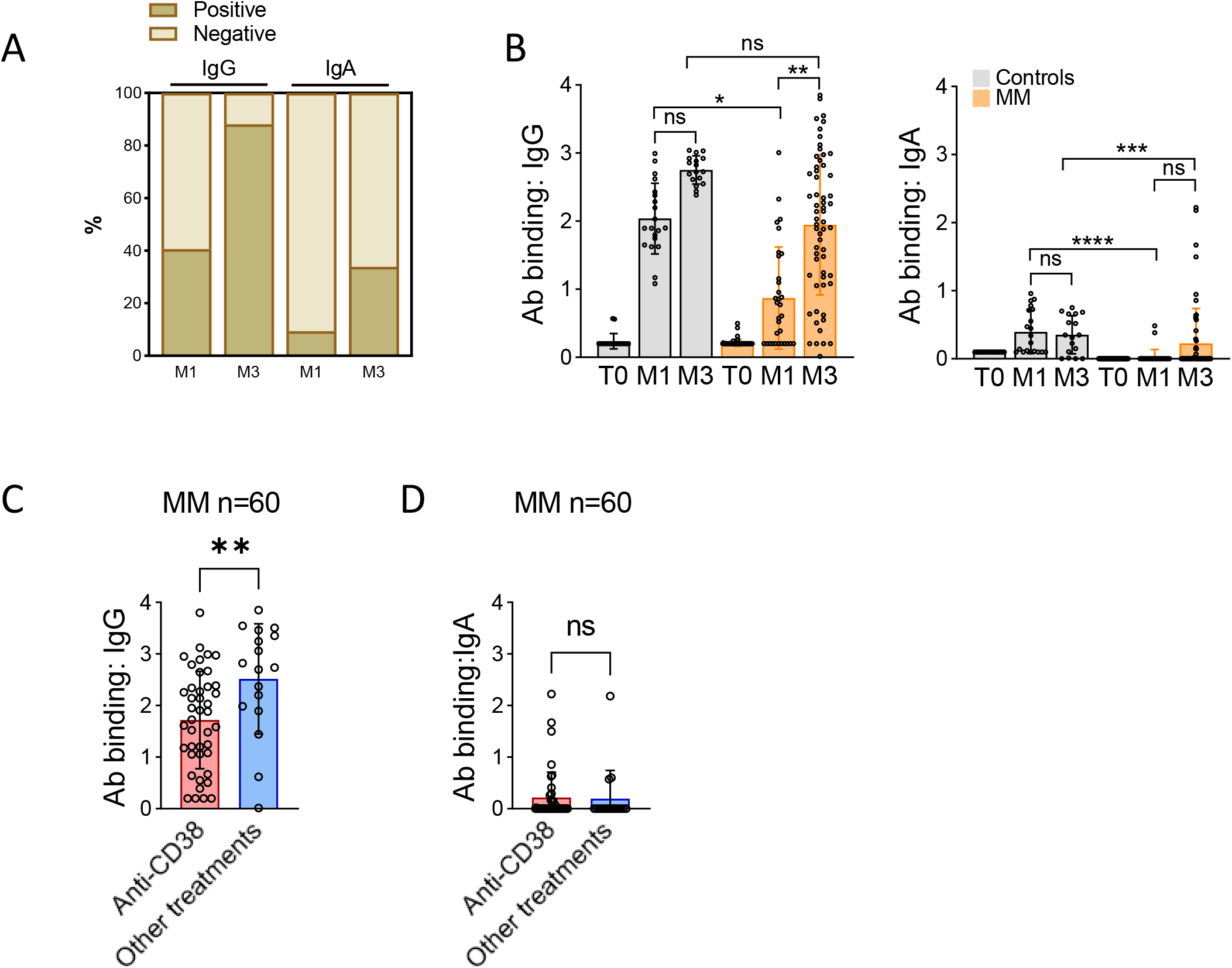
Impaired humoral response to BNT162b2 in multiple myeloma patients. **A-D**. SARS-CoV-2 specific IgG and IgA production quantified by S-flow in 60 and 20 SARS-CoV-2-naive MM patients and healthy volunteers, respectively. **A**. Proportion of IgG and IgA seroconversion with a threshold of 40% and 20%, respectively, at M1 and M3. **B**. IgG (left panel) and IgA (right panel) quantification in controls or MM patients before vaccination (T0) and at M1 and M3. **C-D**. Comparison of IgG (**C**) and IgA (**D**) amounts in MM patients treated or not with anti-CD38 immunotherapy. Histograms represent the mean value. Vertical bars indicate standard error. *p<0.05, **p<0.01, ***p<0.001, ns: not significant.

### Impaired antibody neutralization (NAbs) of alpha and delta variants in myeloma patients

We measured the ability of sera from patients and controls to neutralize alpha and delta variants using the S-Fuse assay. We observed a strong correlation between antibody neutralization against alpha and delta variants, expressed as effective dose 50% (ED50) (**Supplemental Figures 3A-B**). The correlation between SARS-CoV-2 IgG or IgA and alpha or delta NAbs was low, particularly due to patients developing IgG or IgA, but not NAbs (**Figures 3A and Supplemental 3B**). While all subjects in the control group developed NAbs against alpha and delta variants after vaccination, only 51% and 41% of MM patients developed NAbs against alpha and delta variants, respectively (**Figures 3A and Supplemental 3C**). NAbs ED50 was significantly lower in MM patients compared to controls (1747 versus 45, **Figure 3B**), and even lower in MM patients receiving anti-CD38 (**Figure 3C**). The correlation between IgG and NAbs was good in controls (**Supplemental Figure 3D**).

**Figure 3.**
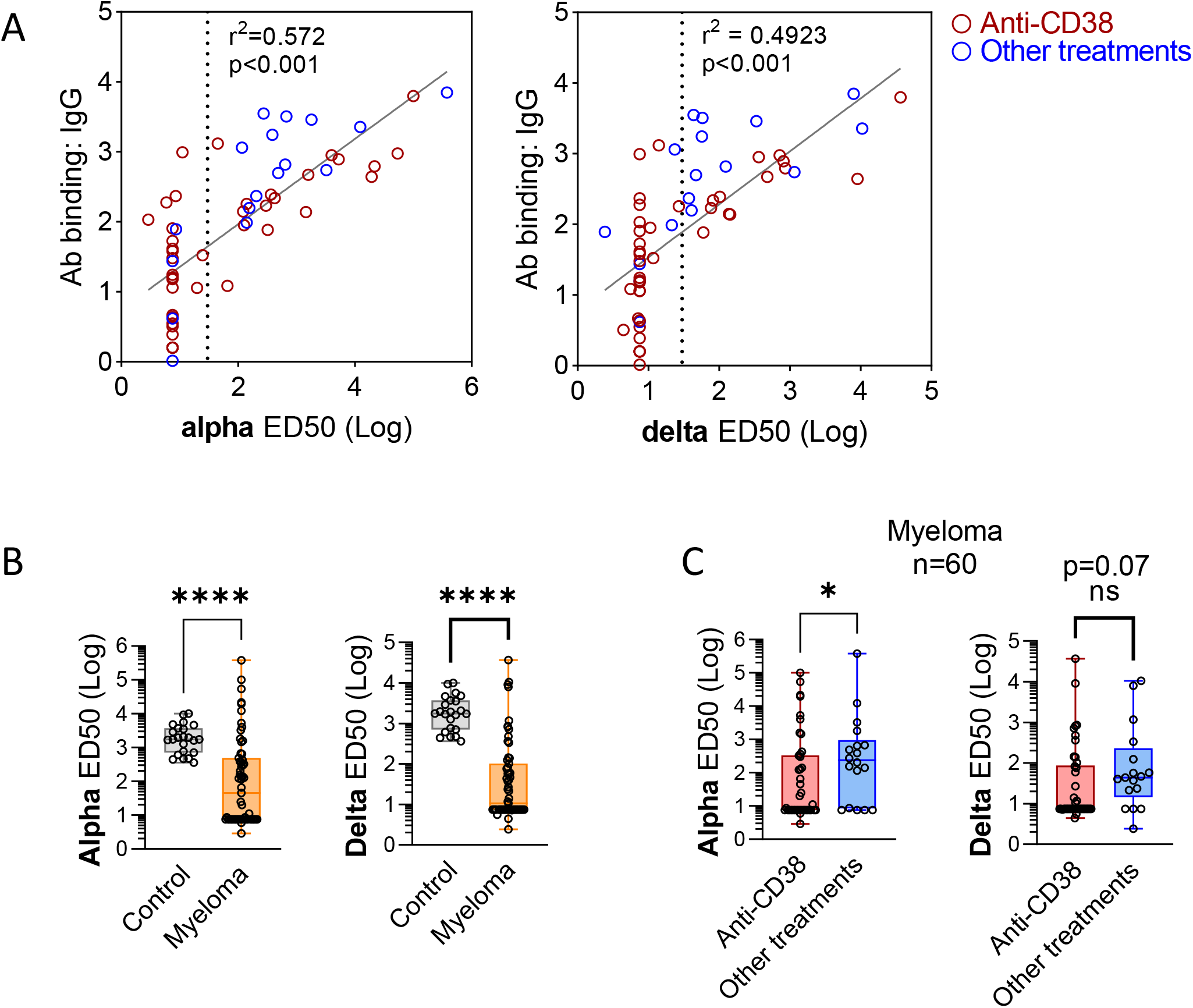
Decreased production of neutralizing antibodies against SARS-CoV-2 alpha and delta variants in myeloma patients. **A**. Linear regression between S-flow IgG and alpha or delta neutralizing antibodies values. Patients receiving anti-CD38 immunotherapies are indicated. The vertical dashed line indicates the threshold for NAbs detection. **B-C**. Quantification of anti-SARS-CoV-2 neutralizing antibodies against alpha or delta variants in controls (n=20) or MM patients (n=60). **B**. Comparison between controls and MM patients. **C**. Comparison between patients receiving or not anti-CD38 immunotherapy. Results are represented as box plots and vertical bars indicate minimum and maximum values. *p<0.05, **p<0.01, ***p<0.001, ns: not significant.

We then investigated factors associated with impaired antibody neutralization in MM patients (**Table 2**). A higher number of treatment lines was significantly associated with a lack of NAbs response, while more patients in first line had a response compared to further lines. Failure to generate a NAbs response was also significantly associated with progressive disease. Among treatment modalities, only anti-CD38 was associated with failure to NAbs response. Lower γ-globulin level was associated with an absence of response, while a lymphocyte count below 0.8×10^9^/l was not. These results show that the post-vaccine production of SARS-CoV-2 NAbs was impaired in MM patients, particularly in case of longer disease evolution and use of anti-CD38 immunotherapy.

**Table 2.** Association between variables and NAbs against alpha or delta variants production. Alpha neutralizing antibodies production was investigated as a binary variable with a cut-off at 30 defining response (more than 30) or absence of response (lower than 30). Ev.: length of MM evolution; y: years; Lines: number of therapeutic lines; ASCT: autologous stem cell transplantation; PD: progressive disease; PI: proteasome inhibitor; Steroids: high-dose steroid therapy ie. more than 20mg dexamethasone per week; γ: corrected gammaglobulin levels; Ig: immunoglobulins; Ly: lymphocytes.

| Alpha ED50 | Response (n=30) | no response (n=30) | p-val | Delta ED50 | Response (n=24) | No response (n=36) | p-val |
| --- | --- | --- | --- | --- | --- | --- | --- |
| Age, y, (min-max) | 68 (40.8-91.7) | 73.7 (49.3-92.1) | 0.07 | Age, y, (min-max) | 68.4 (40.8-91.7) | 72.7 (49.3-92.1) | 0.11 |
| **Disease characteristics** |  |  |  | **Disease characteristics** |  |  |  |
| Ev., y, (min-max) | 4 (0-17) | 5.5 (0-19) | 0.4 | Ev., y, (min-max) | 5.75 (1-17) | 6.34 (0-19) | 0.72 |
| Lines, (min-max) | 1 (1-12) | 2.5 (1-7) | **0.0002** | Lines, (min-max) | 2 (1-12) | 2.85 (1-7) | **0.0004** |
| 1<sup>st</sup> line, n (%) | 17 (55%) | 2 (6.6%) | **0.0001** | 1<sup>st</sup> line, n (%) | 15 (62.5%) | 4 (11%) | **0.0001** |
| ASCT, n (%) | 7 (22.5%) | 1 (3.5%) | 0.05 | ASCT, n (%) | 7 (29%) | 1 (2.7%) | **0.017** |
| PD, n (%) | 4 (12.9%) | 12 (40%) | **0.015** | PD, n (%) | 2 (8.3%) | 13 (36%) | **0.005** |
| **Therapy status** |  |  |  | **Therapy status** |  |  |  |
| Anti-CD38, n (%) | 17 (49%) | 25 (83.3%) | **0.04** | Anti-CD38, n (%) | 13 (54%) | 29 (80.5%) | **0.044** |
| IMiDs, n (%) | 13 (43.3%) | 18 (60%) | 0.19 | IMiDs, n (%) | 10 (41.6%) | 21 (58.3%) | 0.29 |
| PI, n (%) | 7 (23.3%) | 4 (13.3%) | 0.5 | PI, n (%) | 6 (25%) | 5 (13.8%) | 0.32 |
| Steroids, n (%) | 10 (33%) | 12 (40%) | 0.78 | Steroids, n (%) | 9 (37.5%) | 13 (36%) | 0.99 |
| **Biological parameters** |  |  |  | **Biological parameters** |  |  |  |
| γ-globulin < 5 g/l, n (%) | 15 (50%) | 23 (76.6%) | 0.059 | γ-globulin < 5 g/l, n (%) | 11 (45.8%) | 27 (75%) | **0.029** |
| Ly < $0.8 \times 10^9$/l, n (%) | 9 (30%) | 12 (41.4%) | 0.58 | Ly < $0.8 \times 10^9$/l, n (%) | 5 (20.8%) | 16 (44.5%) | 0.096 |

### Reduced cellular immunity in multiple myeloma patients following BNT162b2 vaccination

We performed an IFNγ EliSpot to assess T-cell response upon stimulation with Spike SARS-CoV-2 antigens S1 and S2 in 32 patients of our cohort, including 28 from the anti-CD38 group and 4 from the non-anti-CD38 group. Among them, 26 were SARS-CoV-2 naive patients. The IFNγ response to S1 and S2 peptides was significantly lower in MM patients compared to healthy SARS-CoV-2-naive controls (**Figure 4A**). Numbers of circulating S1 and S2-specificT cells were correlated (**Supplemental Figure 4A**). We then plotted the results of EliSpot against S1 or S2 peptide pools (S1 EliSpot and S2 EliSpot) with the values of NAbs for alpha and delta variants, and observed a strong correlation between both responses (**Figures 4B and Supplemental 4B**). Among SARS-CoV-2-naive patients, we delineated three situations. First, 12 patients (all from the anti-CD38 group) were negative for both NAbs and EliSpot, while SARS-CoV-2 IgG were detected beyond threshold in 8 cases (**Figure 4C**). Secondly, 5 patients who developed a T-cell response against S1 and/or S2 antigen also developed NAbs against alpha and delta variants, and IgG/IgA specific antibodies **(Figure 4C**). Results were discordant in 9 patients, including 3 with positive NAbs without T-cell response, and 6 with the opposite pattern (**Figure 4C**). These results show an impaired T-cell response against SARS-CoV-2 after BNT162b2 vaccine in MM patients, generally associated with an impaired humoral response.

**Figure 4.**
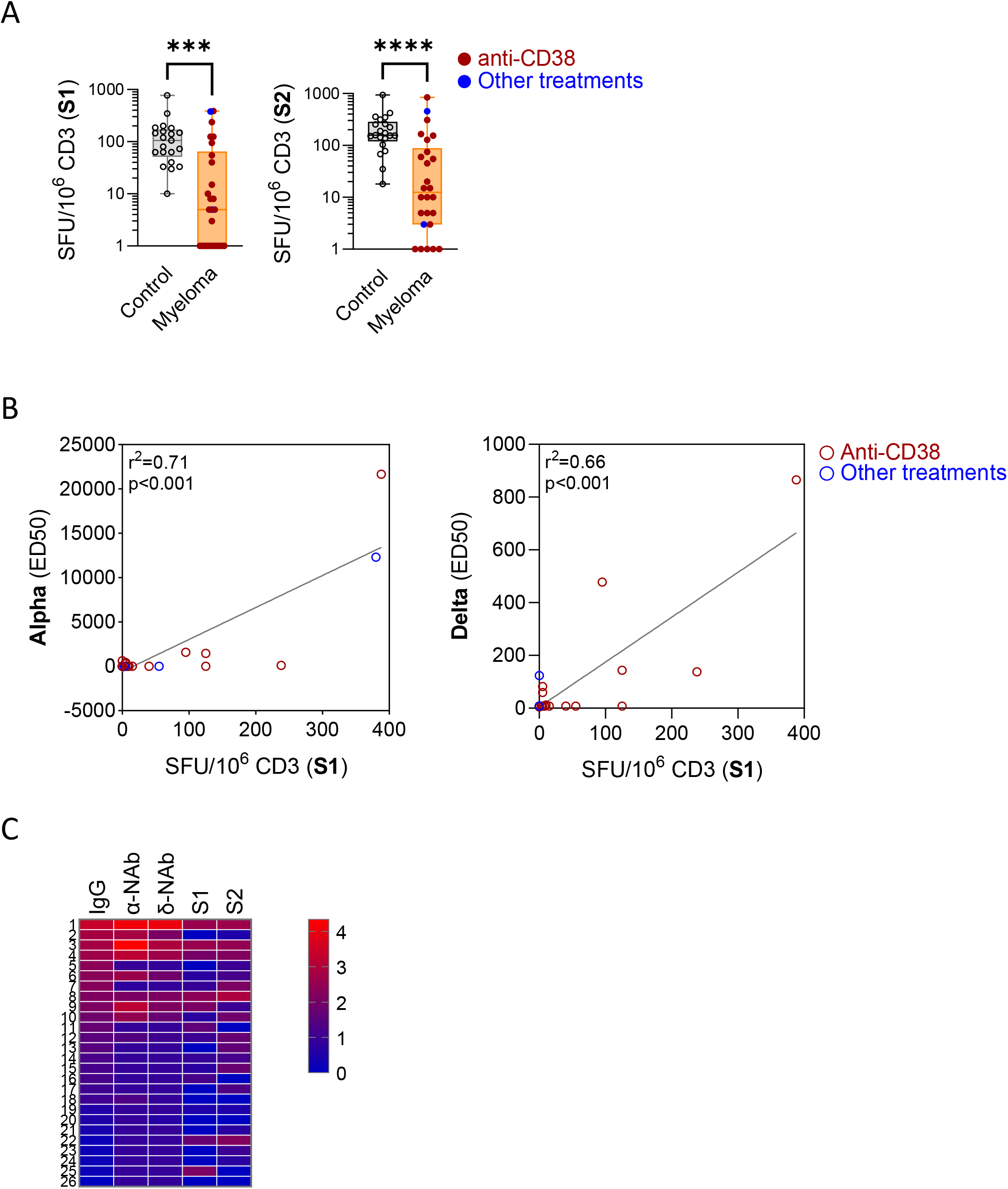
Reduced cellular immunity following BNT162b2 vaccination in multiple myeloma patients. **A**. Quantification of cellular immune response by S1 or S2 EliSpot (in SFU/10^6^ CD3) in controls (n=20) or SARS-CoV-2-naive MM patients (n=26). Patients receiving anti-CD38 immunotherapies are indicated. Results are represented as box plots and vertical bars indicate minimum and maximum values. ***p<0.001. **B**. Linear regression of alpha (left panel) or delta (right panel) neutralizing antibodies values plotted on IFNγ production after exposure to SARS-CoV-2 S1 protein (expressed in SFU/10^6^ CD3). Patients receiving anti-CD38 immunotherapies are indicated. **C**. Heat map representation of IgG S-flow, alpha and delta NAbs and S1 and S2 EliSpot values in 26 SARS-CoV-2-naive MM patients. Results for IgG are linear and NAbs and EliSpot values are log-transformed to facilitate data representation. A color scale is provided.

### Impact of previous SARS-CoV-2 infection on immunological response to BNT162b2 in multiple myeloma patients

We compared the response to BNT162b2 administration in MM patients with or without clinical SARS-CoV-2 infection history. Among the 72 MM patients included in our study, 11 were previously exposed to SARS-CoV-2, including six and five in the anti-CD38 and non-anti-CD38 groups, respectively. We measured anti-SARS-CoV-2 specific IgG as well as alpha and delta NAbs (n=11), and performed EliSpot assays (n=5) in these patients at M3. We compared these results with those of MM patients after excluding subject who suffered from SARS-CoV-2 infection after vaccination. IgG levels were significantly higher in MM patients with a history of SARS-CoV-2 infection (before or after vaccination) compared to those without, independently from the timing of vaccination (**Figure 5A**). Similar results were observed for alpha and delta specific NAbs levels (**Figure 5B**). Moreover, while based on a limited number of patients, we observed a similar profile concerning S1 and S2 EliSpot assays (**Figure 5C**). Collectively, these results suggest that the immune response achieved after BNT162b2 vaccine was lower compared to that achieved after SARS-CoV-2 infection in immunocompromised MM patients.

**Figure 5.**
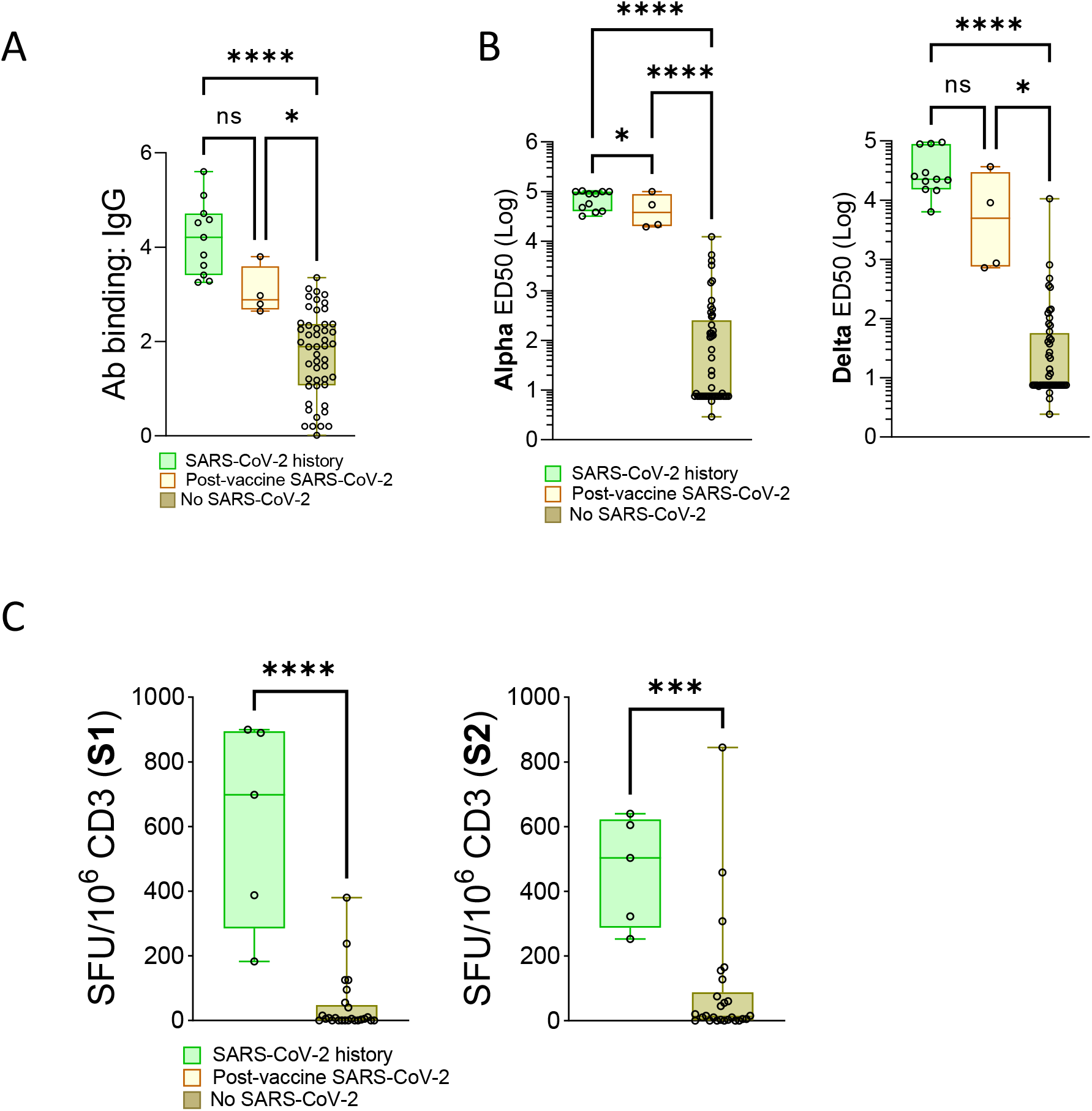
Impact of previous SARS-CoV-2 infection on immunological response to BNT162b2 in multiple myeloma patients. **A-C**. MM patients with SARS-CoV-2 history (more than three month before vaccination), having developed SARS-CoV-2 infection at any time after vaccination or SARS-CoV-2 naive. **A**. S-flow IgG quantification. **B**. Alpha (left) or delta (right) NAbs quantification. **C**. S1 (left) or S2 (right) EliSpot. Results are represented as box plots and vertical bars indicate minimum and maximum values. *p<0.05, **p<0.01, ***p<0.001, ns: not significant.

### Clinical response to to BNT162b2 in MM patients

We did not observe any clinical recurrence in the 11 patients with previously documented SARS-CoV-2 infection. Among the 61 patients without previously documented SARS-CoV-2 infection, four developed an infection and belonged to the anti-CD38 group (**Figure 1**). SARS-CoV-2 infection occurred 9 and 19 days after the first vaccine dose in two patients, and 16 and 23 days after the second dose in two others. These patients required hospitalization; half of them received conventional oxygen therapy, but none required intensive care. They all fully recovered and cleared SARS-CoV-2 as determined by negative PCR testing on nasopharyngeal swab.

We also investigated SARS-CoV-2-related death that occurred in all 39 AP-HP hospitals from January 2020 to July 2021. We focused on two epidemic peak periods before (period 1, March-July 2020) and after (period 2, March-July 2021) vaccination program onset. We observed 2764 and 1842 deaths overall, and 42 and 39 deaths in MM patients during period 1 and period 2, respectively (**Supplemental Figures 5A-B**). While the prescription of anti-CD38 therapies was stable at these AP-HP centres between period 1 and period 2 (mean cycle per month 1618 and 1841 during period 1 and period 2, respectively, p=0.15, **Supplemental Figure 1C**), we observed that the proportion of death among MM patients receiving anti-CD38 was stable (11 and 17 death during period 1 and period 2, respectively), while the number of death among patients not receiving anti-CD38 tended to decrease (31 and 22 death during period 1 and period 2, respectively) (**Table 3**). Altogether, these results suggest that anti-CD38 therapy could decrease the efficacy of mRNA SARS-CoV-2 vaccine in MM patients.

**Table 3.** Evolution of SARS-CoV-2-related mortality in AP-HP healthcare centres between 2020 and 2021. 2020: March to July 2020; 2021: March to July 2021. General: total number of SARS-CoV-2-related death regardless underlying conditions.

|  | **Anti-CD38** | **Other treatment** | **Myeloma (total)** | **General** | **Anti-CD38 cycles (n)§** |
| --- | --- | --- | --- | --- | --- |
| 2020* | 11 | 31 | 42 | 2764 | 8091 |
| 2021* | 17 | 22 | 39 | 1942 | 9206 |
| Difference | +54% | -29% | -7% | -33% | +13.5% |

## Discussion

Multiple myeloma was initially considered as a high-priority condition for vaccination against SARS-CoV-2 (ESMO, no date). However, some uncertainties exist regarding the immune response to vaccine, especially in patients receiving anti-CD38 immunotherapy and against emerging variants of concern. Since high titers of neutralizing antibodies are necessary to inhibit delta variant (Planas, Veyer, *et al*., 2021), patients with impaired immune response may be at risk of breakthrough infection, despite completing full vaccination. In our study, we prospectively investigated the humoral and cellular responses after 2 doses of BNT162b2 in a cohort of MM patients.

According to standard procedures, we investigated humoral response with the S-flow method, evaluating the binding of SARS-CoV-2 specific IgG and IgA to Spike-expressing cells (Grzelak *et al*., 2020). The proportion of MM patients achieving humoral response was modest after the first injection (44% and 9% for IgG and IgA, respectively), but largely increased after the second dose (85% and 34% for IgG and IgA, respectively). These results contrast with the robust immune response observed in vaccine trials done in healthy subjects (Polack *et al*., 2020; Baden *et al*., 2021), but were consistent with previous reports on patients with haematological malignancies. Notably, Pimpenelli and colleagues studied the immunogenicity of anti-SARS-CoV-2 BNT162b2 vaccine in MM, and showed a serocoversion rate of 21% and 78.6% after the first (week 3) and second dose (week 5), respectively (Pimpinelli *et al*., 2021). These results emphasize the requirement of a whole vaccination procedure before evaluating the response to vaccination (Boyarsky *et al*., 2021). Moreover, IgG responses were heterogeneous in MM patients compared to healthy controls, as reported (Van Oekelen *et al*., 2021).

To better assess the protection conferred by BNT162b2 in our cohort of MM patients, we investigated antibody neutralization (NAbs) against SARS-CoV-2 variants of concern. The presence of NAbs is highly predictive of protection against coronavirus infection in healthy subjects (Barnes *et al*., 2020; Rogers *et al*., 2020; Khoury *et al*., 2021). We investigated NAbs against the two variants predominantly found in the French population at the time of our data collection, namely alpha and delta. We observed that MM patients had a significantly lower NAbs response compared to healthy subjects, and that anti-CD38 therapy correlated with an even lower NAbs response among MM patients. Our results suggested that the correlation between S-flow and alpha and delta NAbs values was not as robust in MM patients as in healthy controls, especially in the case of anti-CD38 immunotherapy. Thus, a positive serology has most likely a low predictive value of in terms of protection against SARS-CoV-2 infection in case of sub-optimal responses.

We also enumerated the number of circulating Spike SARS-CoV-2-specific IFNγ-producing T-cells using an EliSpot assay (Nelde *et al*., 2021). The induction of a T-cell response correlated with the levels of anti-SARS-CoV-2 NAbs. In contrast to healthy subjects, some MM patients showed discrepancies between humoral and cellular responses to BNT162b2, reflecting the profound impairment of their immunoglobulin production capacities. It was reported in vaccine trials that the BNT162b2 induced a specific T-cells response in healthy individuals (Walsh *et al*., 2020). On the contrary, in MM patents, T cell responses are weaker compared to healthy controls and highly heterogenous.

Immunotherapy with monoclonal anti-CD38 antibodies recently emerged as a paradigm shift in the management of MM, with significant benefits in survival (van de Donk, Richardson and Malavasi, 2018). Isatuximab or Daratumumab, in association with dexamethasone and/or chemotherapy, frequently induce profound and durable response including in heavily pre-treated MM patients (Attal *et al*., 2019; Bahlis *et al*., 2020; Dimopoulos *et al*., 2020). Unfortunately, these highly active monoclonal antibodies also reduce the frequency of normal bone marrow plasma cell in MM patients. Anti-CD38 antibodies also induce a partial natural killer (NK)-cells depletion, which could contribute to immunodeficiency in MM receiving these immunotherapies (Nahi *et al*., 2019). Accordingly, we observed a significant reduction of polyclonal IgG, but not of lymphocyte count, in MM patients treated with anti-CD38 compared to patients receiving alternative therapies.

From a clinical perspective, epidemiological data from AP-HP healthcare centres showed that SARS-CoV-2-related mortality among MM patients between March-July 2020 and 2021 decreased, similar to that of the general population, except in patients treated with anti-CD38, although the prescriptions of these immunotherapies remained stable across these periods. Moreover, we observed a clinical SARS-CoV-2 infection in four MM patients after vaccination, all actively treated with anti-CD38 immunotherapies. While currently not statistically significant, this difference could represent a warning and incite to an even closer monitoring of SARS-CoV-2-related symptoms in patients receiving such treatments.

MM patients with prior SARS-CoV-2 infection demonstrated a homogeneous immune response after vaccination. They presented robust T-cell response and significantly higher NAbs titer than in SARS-CoV-2-naive patients. Vaccination after SARS-CoV-2 infection results in a potent immune response in the healthy population (Stamatatos, *et al*., 2021; Planas, Veyer, *et al*., 2021). These findings shows that MM patients mount a good immune response in certain circumstances, strongly suggesting that alternative vaccination strategies may elicit an efficient protection against SARS-CoV-2. The use of booster strategies, such as two doses of Influenza vaccine or administration of conjugate vaccines for *Streptococcus pneumonia* infections are currently used invesitgation in patients with haematological malignancies (Ludwig *et al*., 2021). The administration of a third dose of vaccine could improve immune response and NAbs production, particularly in the case of treatment with anti-CD38 immunotherapy.

Our study had several limitations. Our main readout of anti-SARS-CoV-2 vaccine response was the production of NAbs, for which the threshold needed to protect patients from SARS-CoV-2 is still unknown. We investigated T-cell response in only half of the patients, due to technical limitations. The follow-up to evaluate SARS-CoV-2 infection incidence and severity was limited in time. Follow-up studies of larger cohorts of MM having received SARS-CoV-2 vaccine are needed to clearly determine the impact of vaccination strategies in this severely immunocompromised population.

To summarize our findings, vaccinated MM patients had a delayed seroconversion, a decreased NAbs production, and an impaired cellular response to SARS-CoV-2 compared to healthy control the future evaluation of alternative strategies such as booster protocols will help defining an adapted procedure for MM patients and other severely immunocompromised patients.

## Data Availability

The data that support the findings of this study are available from the corresponding author, upon reasonable request.

## Acknowledgments

The authors first thank the patients and the healthy volunteers who participated in this study. We are very grateful to all nurses, technicians and physicians from Groupe Hospitalo-Universitaire AP-HP.Centre-Université de Paris involved in the daily care of the patients included or not in this study, who have gone far beyond their usual jobs during this epidemic.

## Author contributions

SH, JZ, TB, AO and DP designed the study, collected and analyzed data and wrote the manuscript ; TB, AO, DP and IS designed and performed experiments ; PD, JH, BV, FS, NE, DB, LW, GF, JD, PF and BD took care of the patients ; BT and JT analyzed data and wrote the manuscript ; LC and OS designed and supervised experiments and wrote the manuscript ; MV designed and supervised the study, analyzed data and worte the manuscript.

---

**Supplemental Figure 1.**
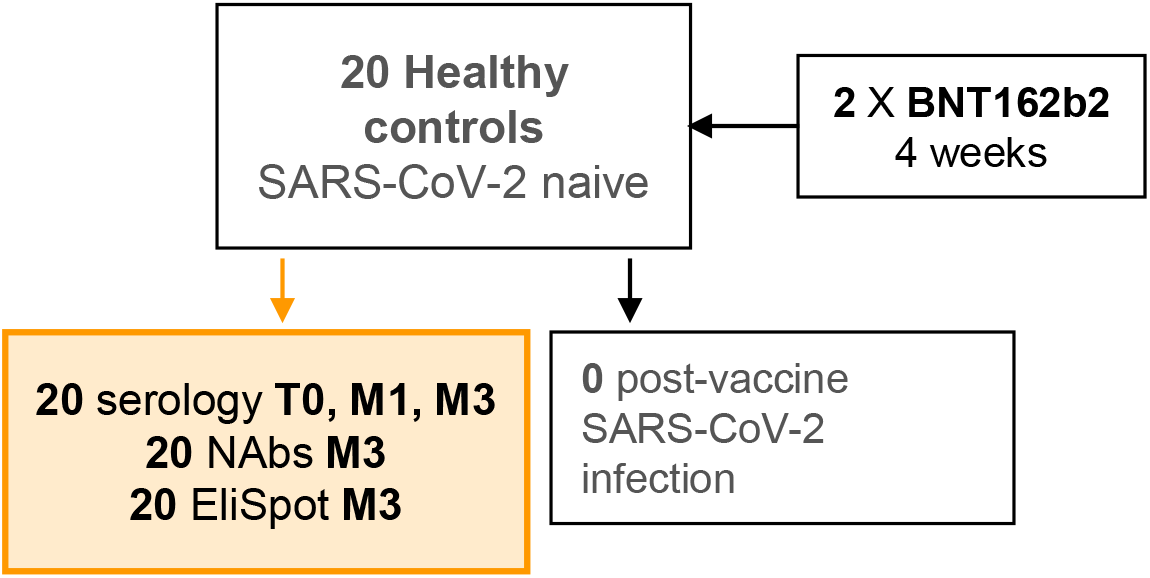
Flow chart of healthy controls. Flow-chart of healthy controls prospectively included in our study. 2 X BNT162b2: two doses of BNT162b2 mRNA vaccine. Serology: specific anti-SARS-CoV-2 IgG or IgA quantification by the S-flow method; NAbs: neutralizing antibodies against alpha or delta SARS-CoV-2 variants; EliSpot: IFNγ production in response to S1 or S2 SARS-CoV-2 proteins. M1 and M3: one and three months after the first dose of BNT162b2.

**Supplemental Figure 2.**
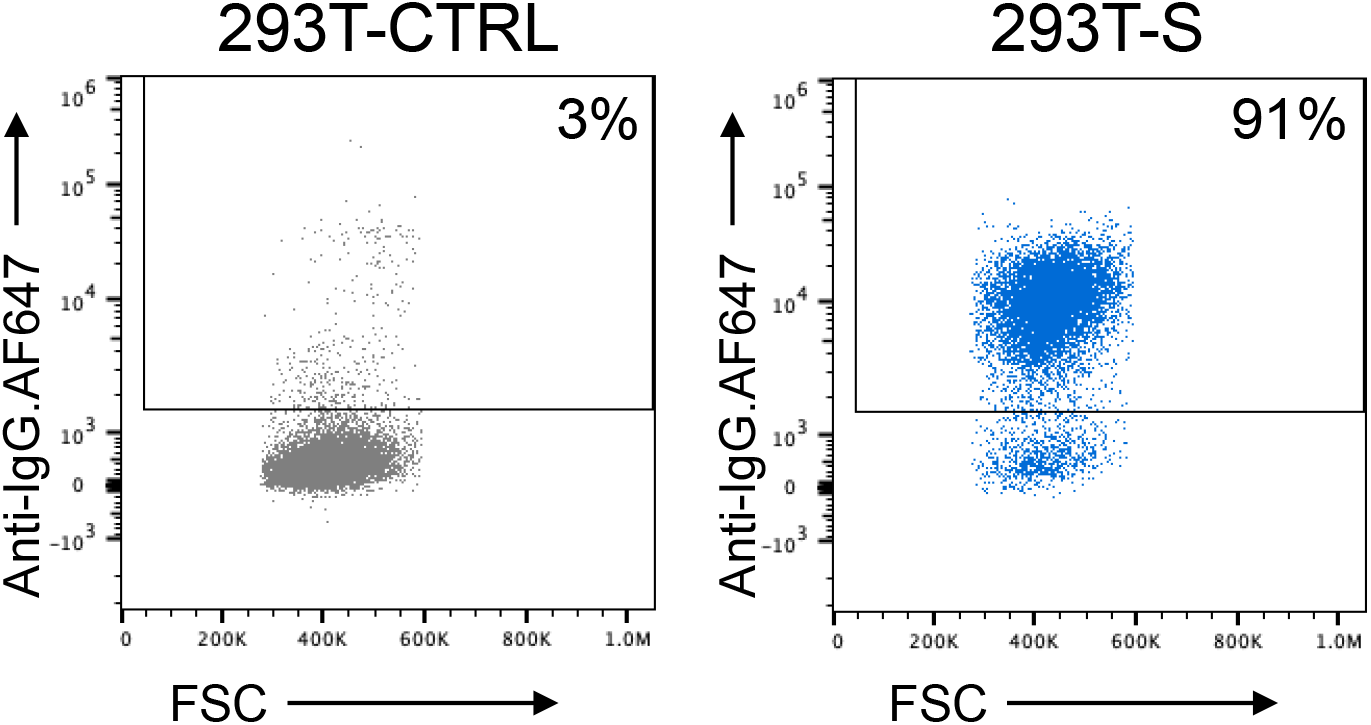
S-Flow assay. Recognition of SARS-CoV-2 S-protein expressed on the surface of 293T cells (293T-S) using flow cytometry (anti-spike IgG). 293T-CTRL: 293T Empty cells. 293T-S: 293T cells stably expressing the S protein (293T Spike cells).

**Supplemental Figure 3.**
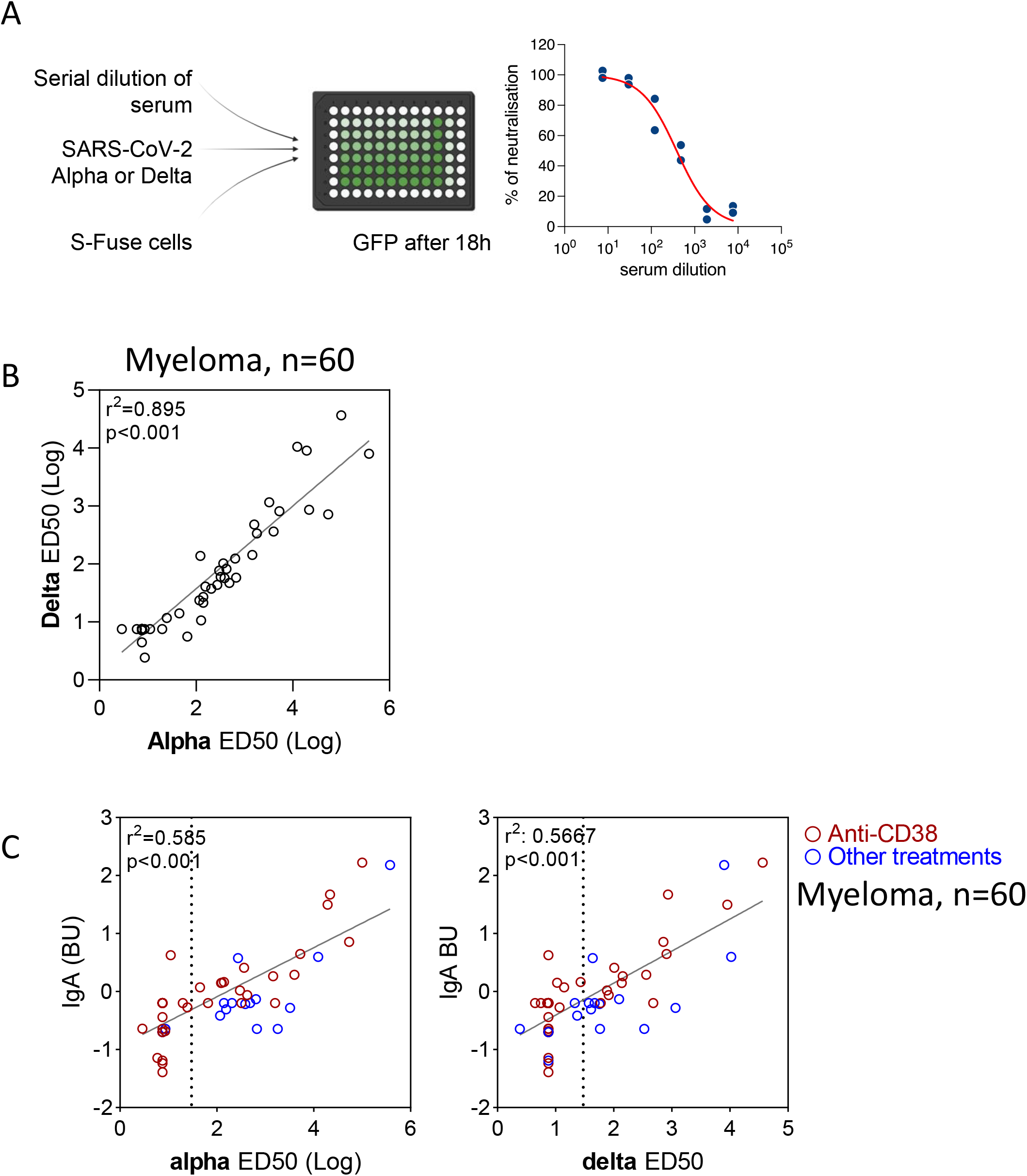
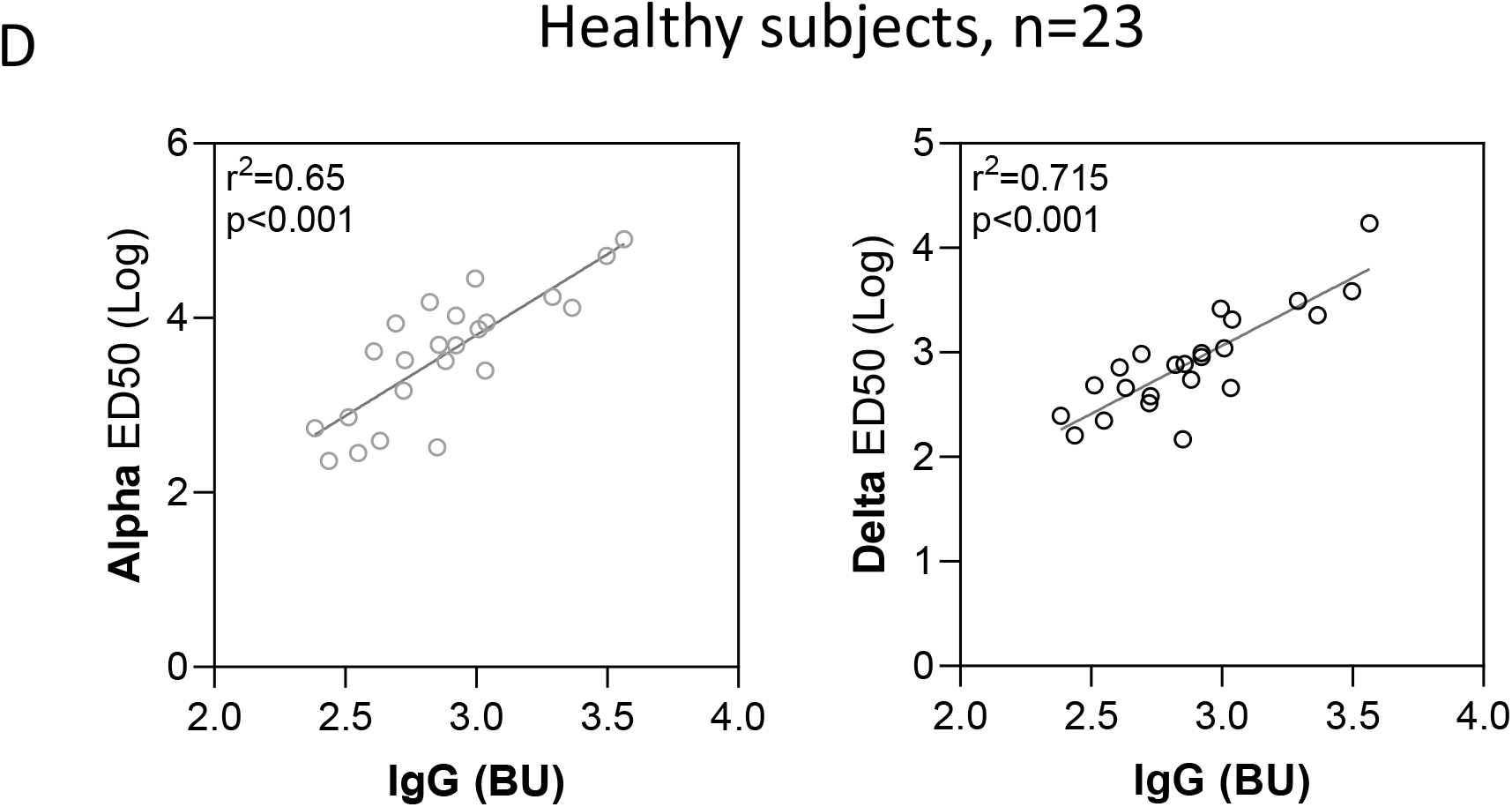
Production of neutralizing antibodies against SARS-CoV-2 alpha and delta variants. **A**. S-Fuse assay. **B**. Linear regression between antibody neutralization against alpha and delta variants, expressed as median effective dose (ED50), in MM. **C**. Linear regression between SARS-CoV-2 specific IgA and alpha or delta neutralizing antibodies values, in MM. Patients receiving anti-CD38 immunotherapies are indicated. The vertical dashed line indicates the threshold for NAbs detection. **D**. Linear regression between antibody neutralization against alpha or delta variants, and SARS-CoV-2 specific IgG in healthy subjects.

**Supplemental Figure 4.**
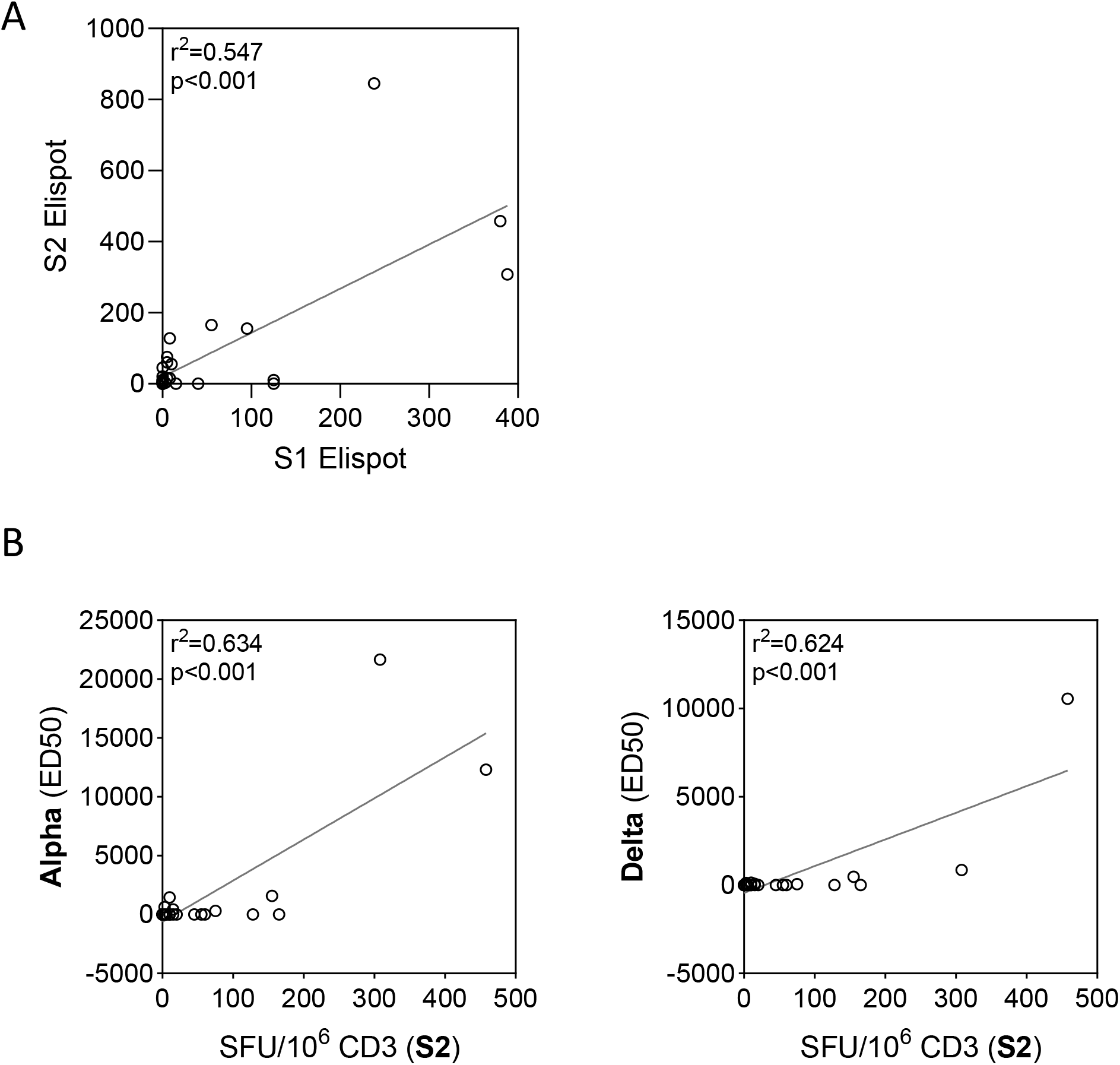
Correlation IFNγ production and NAbs after full BNT162b2 in MM. **A.**Linear regression between EliSpot against S1 or S2 antigen (S1 EliSpot and S2 EliSpot). **B**. Linear regression of alpha (left panel) or delta (right panel) neutralizing antibodies values plotted on IFNγ production after exposure to SARS-CoV-2 S1 protein (expressed in SFU/10^6^ CD3).

**Supplemental Figure 5.**
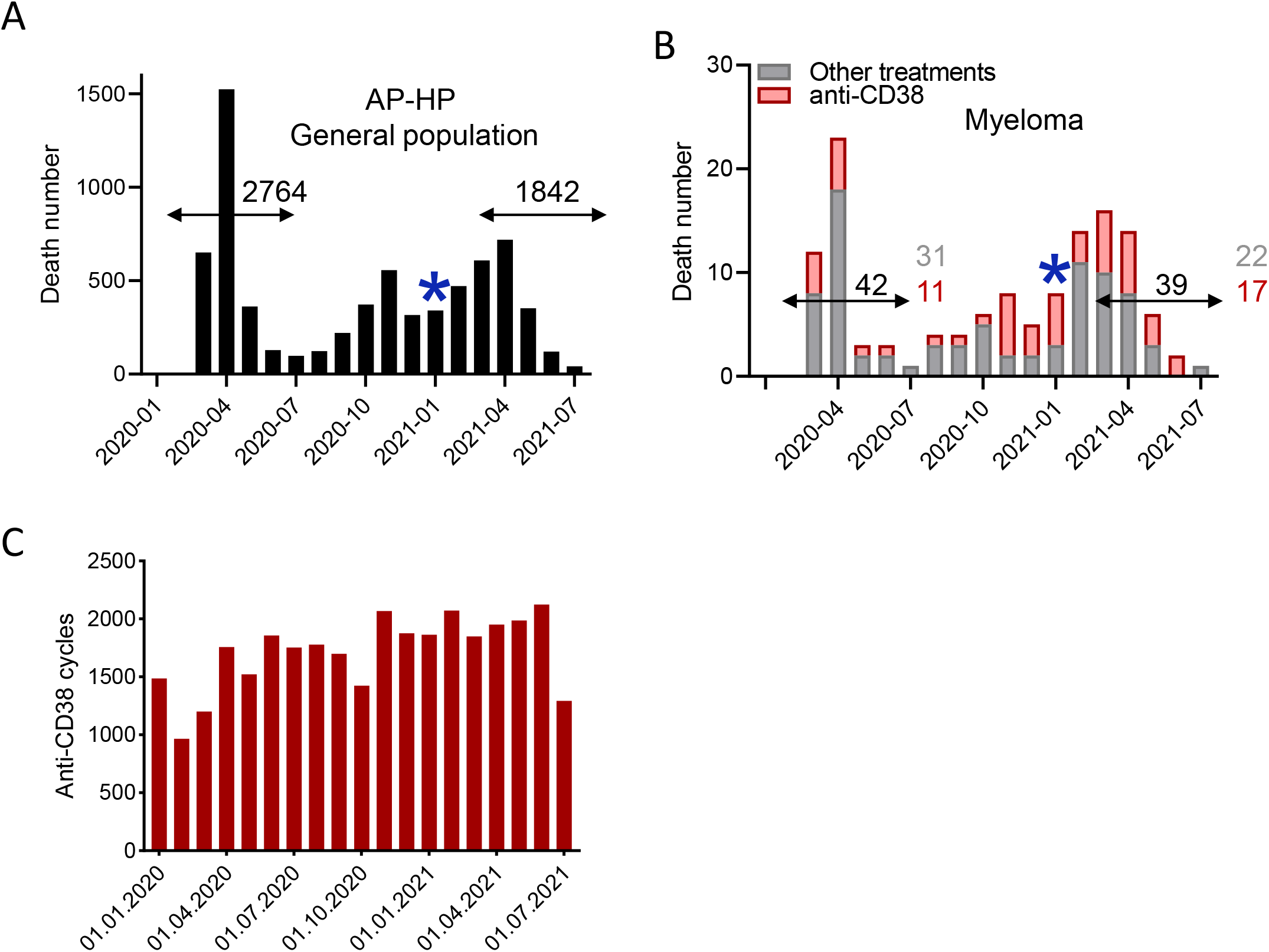
Clinical response to BNT162b2 in healthy controls and SARS-CoV-2-related death before and after vaccination program onset. **A-B**. SARS-CoV-2 related death in AP-HP hospitals before (period 1, March-July 2020) and after (period 2, March-July 2021) vaccination program onset. The star corresponds to the beginning of the vaccination program. **A**. In general population. **B**. In MM (patients receiving anti-CD38 and others). **C**. Evolution of the prescription of anti-CD38 therapies evolution at AP-HP centres.

